# Plasma Proteomic Mediators of the Association Between Early-Pregnancy Psychosocial Stress and Cardiovascular Health 2 to 7 Years After Delivery: The nuMoM2b-HHS Study

**DOI:** 10.64898/2026.09.14.26363041

**Authors:** Xiaoning Huang, Lucia C. Petito, Weihua Guan, Lynn M. Yee, David M. Haas, Brian M. Mercer, Samuel Parry, George R. Saade, Robert M. Silver, Yi Qiao, Xiaomeng Huang, Hyagriv N. Simhan, Uma M. Reddy, Judith Chung, Philip Greenland, Donald M. Lloyd-Jones, William A. Grobman, Sadiya S. Khan

**Affiliations:** Northwestern University Feinberg School of Medicine, Department of Medicine, Division of Cardiology, Chicago, IL; Northwestern University Feinberg School of Medicine, Department of Preventive Medicine, Chicago, IL; University of Minnesota, School of Public Health, Division of Biostatistics & Health Data Science; Northwestern University Feinberg School of Medicine, Department of Obstetrics and Gynecology, Division of Maternal-Fetal Medicine, Chicago, IL; Indiana University School of Medicine, Department of Obstetrics and Gynecology, Indianapolis, IN; Case Western Reserve University School of Medicine, Department of Obstetrics and Gynecology, Cleveland, OH; University of Pennsylvania School of Medicine, Department of Obstetrics and Gynecology, Division of Maternal-Fetal Medicine, Philadelphia, PA; University of Texas Medical Branch, Department of Obstetrics and Gynecology, Galveston, TX; University of Utah Health Sciences Center, Department of Obstetrics and Gynecology, Salt Lake City, UT; Utah Center for Genetics Discovery, University of Utah School of Medicine, Salt Lake City, UT; University of Pittsburgh School of Medicine, Magee-Women’s Research Institute & Foundation, Department of Obstetrics and Gynecology, Pittsburgh, PA; Columbia University Irving Medical Center, Department of Obstetrics and Gynecology, New York, NY; University of California Irvine School of Medicine, Department of Obstetrics and Gynecology, Division of Maternal-Fetal Medicine, Irvine, CA; Preventive Medicine & Epidemiology, Boston University Chobanian & Avedisian School of Medicine, Boston, MA; The Warren Alpert Medical School of Brown University, Department of Obstetrics and Gynecology, Division of Maternal-Fetal Medicine, Providence, RI

## Abstract

**Importance:** Psychosocial exposures in early pregnancy are associated with worse maternal cardiovascular health (CVH), but the biological pathways linking early-pregnancy stressors to long-term CVH are not defined.

**Objective:** To determine whether plasma proteins measured in early pregnancy mediate the association between early-pregnancy psychosocial stressors and CVH 2 to 7 years after delivery.

**Design, Setting, and Participants:** This was a secondary analysis of the prospective multicenter Nulliparous Pregnancy Outcomes Study: Monitoring Mothers-to-Be Heart Health Study (nuMoM2b-HHS), which enrolled nulliparous individuals at eight US academic centers (2010-2014) and followed them 2 to 7 years after delivery. The analytic sample comprised participants with first-trimester plasma proteomics. Analyses were conducted from January to June 2026.

**Exposures:** Perceived stress (Perceived Stress Scale-10 >13) as the primary exposure; Early-pregnancy depression (Edinburgh Postnatal Depression Scale >10) and anxiety (State-Trait Anxiety Inventory >37) as secondary exposures.

**Main Outcomes and Measures:** Post-pregnancy CVH quantified by the American Heart Association Life’s Essential 8 (LE8) composite score (0-100) and its clinical components: blood pressure, lipid, glucose, and body mass index (BMI) scores. A four-way decomposition partitioned the total effect of stress into a controlled direct effect, reference interaction, mediated interaction, and pure indirect effect operating through a proteomic mediator.

**Results:** Among 1662 participants (mean [SD] age, 27.1 [5.5] years), 41.6% screened positive for stress. Of 6874 aptamers, 17 (representing 14 unique, predominantly neuronal and synaptic proteins) were associated with both stress and the LE8 composite as well as four clinical components after false discovery rate correction. No aptamers met criteria for depression or anxiety. Stress was associated with lower levels of all 14 proteins, and higher protein levels were associated with better CVH scores. The 14 proteins reduced to a single principal component (57% of variance) used as the mediator. Stress was associated with a lower post-pregnancy LE8 composite score (total effect, −3.74 [95% CI, −5.03 to −2.45]); the proteomic factor accounted for a pure indirect effect of −1.20 (95% CI, −1.68 to −0.72), approximately 32% of the total effect, and approximately 56% of the larger association with the BMI score. Significant indirect effects through the proteomic factor were also observed for the blood pressure, lipid, and glucose scores despite nonsignificant total effects.

**Conclusions and Relevance:** A coherent neuronal and synaptic plasma protein signature in early pregnancy mediated a substantial share of the association between maternal psychosocial stress and cardiovascular health years after delivery, identifying a candidate biological pathway for further study.

**Key Points:** *Question:* Do plasma proteins measured in early pregnancy mediate the association between early-pregnancy psychosocial stress and cardiovascular health (CVH) 2 to 7 years after delivery?

*Findings:* In this analysis of 1662 nulliparous individuals, early-pregnancy perceived stress was associated with lower post-pregnancy Life’s Essential 8 CVH. A single factor derived from 14 predominantly neuronal and synaptic plasma proteins mediated approximately one-third of this association and more than half of the association with body mass index.

*Meaning:* A coherent neuronal and synaptic plasma protein signature in early pregnancy may represent a biological pathway linking maternal psychosocial stress to long-term cardiovascular health.

## Introduction

The peripartum period is a critical window for assessing and modifying long-term maternal cardiovascular risk. ^1–3^ The American Heart Association defines cardiovascular health (CVH) using 8 clinical and behavioral factors collectively termed Life’s Essential 8 (LE8) summarized as a composite score from 0 to 100, with higher scores indicating better health. ^4^ Poor CVH in pregnancy is associated with adverse pregnancy outcomes and with cardiovascular disease across the life course, and CVH in the years after a first birth is an important determinant of a woman’s long-term cardiovascular trajectory. ^1,3,5,6^

Psychosocial stress is associated with cardiovascular dysfunction and disease burden in women, and pregnancy is a period of amplified psychosocial stress. ^7^ Elevated perceived stress in pregnancy has been linked to suboptimal CVH, to adverse pregnancy outcomes, and to higher blood pressure in the years after delivery, with effects that may persist well beyond the perinatal period. ^7^ Yet the biological mechanisms that connect early-pregnancy psychosocial stress to CVH measured years later remain poorly understood, limiting opportunities for mechanistic insight and intervention.

High-throughput plasma proteomics offers an agnostic window into the biological pathway that links exposures to downstream cardiometabolic phenotypes. Large-scale aptamer-based proteomics have been applied in early pregnancy in this cohort to evaluate prediction of hypertensive disorders of pregnancy. ^8^ However, to our knowledge, whether specific proteins lie on the pathway between maternal psychosocial stress and long-term CVH and could potentially serve as candidate mediators has not been examined.

We hypothesized that a subset of early-pregnancy plasma proteins would mediate the association between early-pregnancy psychosocial stress and post-pregnancy CVH. Using the nuMoM2b-HHS cohort, we screened the plasma proteomics for proteins jointly associated with early pregnancy psychosocial exposures and post-pregnancy CVH outcomes, reduced the candidate proteins to a parsimonious factor, and quantified mediation using a four-way decomposition that separates mediation from exposure-mediator interaction.

## Methods

### Study Population

This was a secondary analysis of the Nulliparous Pregnancy Outcomes Study: Monitoring Mothers-to-Be (nuMoM2b) and its Heart Health Study (nuMoM2b-HHS) follow-up. The nuMoM2b study enrolled nulliparous individuals with singleton pregnancies and cardiac activity at less than 14 weeks of gestation between 2010 and 2014 across 8 US clinical sites affiliated with tertiary care centers. ^9^ The nuMoM2b-HHS re-examined a subsample 2 to 7 years after delivery to characterize CVH after a first birth. ^9^ Institutional review board approval and written informed consent were obtained at each participating site. The present analysis included nuMoM2b-HHS participants with first-trimester plasma proteomics available (analytic sample, N=1662; **Figure 1**). Characteristics of participants with and without proteomics are compared in **eTable 1** in the Supplement.

**Figure 1.**
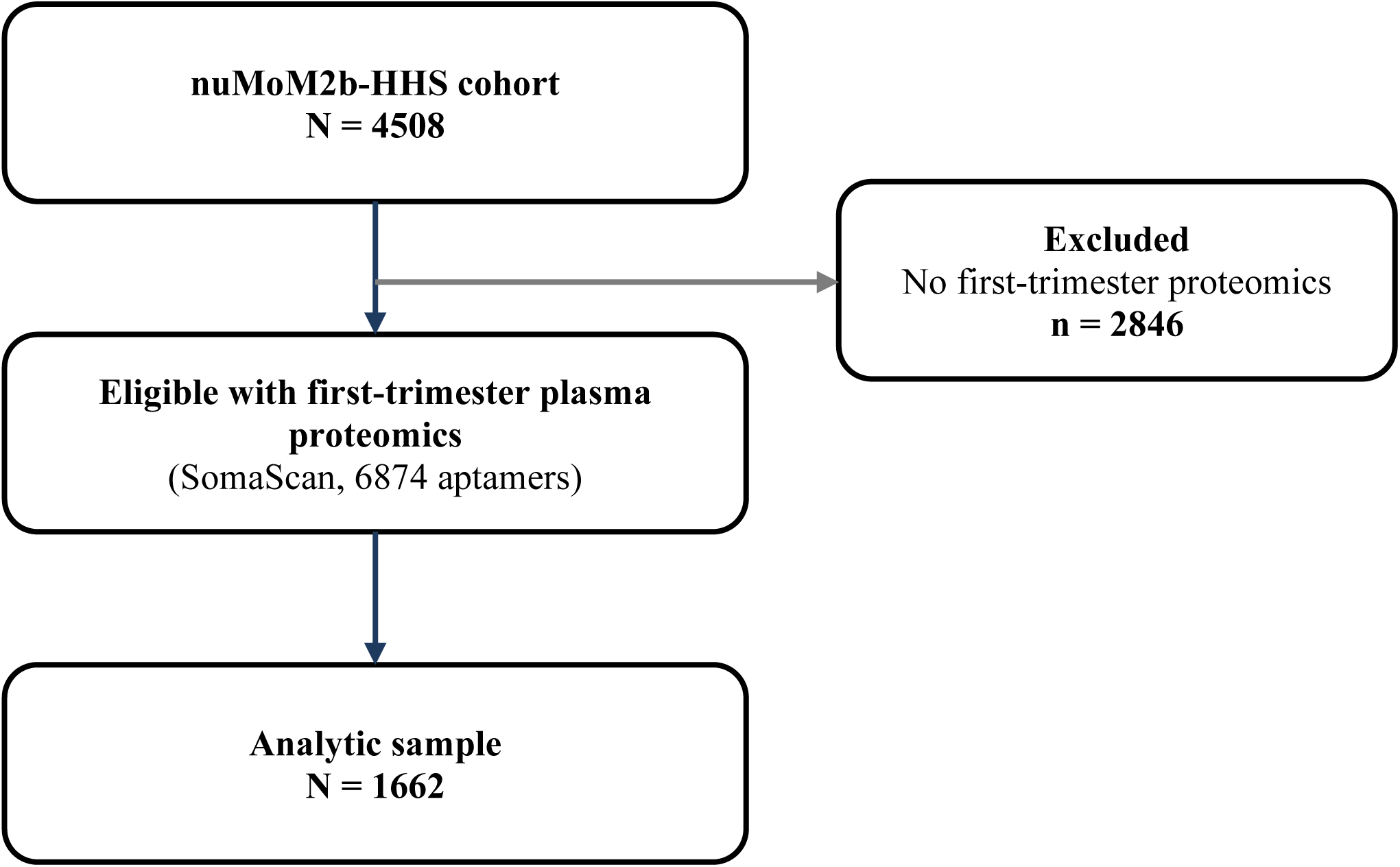
Flow chart for the nuMoM2b-HHS cohort to the analytic sample. Of 4508 nuMoM2b-HHS participants, 2846 lacked first-trimester proteomics, yielding an analytic sample of 1662 participants with proteomic, psychosocial, and post-pregnancy cardiovascular health data.

### Psychosocial Exposures

Psychosocial stress and depression were measured by questionnaires in early pregnancy. Anxiety was measured in mid-pregnancy and was used as proxy to anxiety levels in early pregnancy when plasma proteomics were collected. Perceived stress was measured with the 10-item Perceived Stress Scale (PSS-10; range, 0-40) and dichotomized as stress at a score greater than 13. ^10^ Depressive symptoms were measured with the Edinburgh Postnatal Depression Scale (EPDS; range, 0-30) and dichotomized at 10 or higher for depressive symptoms. ^11,12^ Anxiety was measured with the State-Trait Anxiety Inventory (STAI; range, 0-60) and dichotomized at greater than 20. ^13,14^ All three exposures were evaluated as candidate exposures. Stress was the primary exposure carried into mediation analysis based on the screening results described below.

### Plasma Proteomics

First-trimester plasma was profiled using a modified aptamer-based assay (SomaScan) that quantified 6874 aptamers targeting unique human proteins. ^15^ Following standard quality control, protein values were log2-transformed and standardized to a mean of 0 and SD of 1 prior to analysis.

### Cardiovascular Health Outcomes

Post-pregnancy CVH at the nuMoM2b-HHS visit (2-7 years after delivery) was quantified using the LE8 framework. ^4^ Each component was scored 0 to 100, and the composite LE8 score was the mean of the eight component scores. The prespecified primary outcome was the composite LE8 score. Because proteins are mechanistically most plausible as mediators of the cardiometabolic components, the four clinical component scores, including blood pressure, blood lipids, blood glucose, and BMI, were analyzed as secondary outcomes.

### Statistical Analysis

Missing covariate and outcome data were handled with multiple imputation by chained equations (10 imputed datasets). ^16^ Estimates were combined across imputations using Rubin’s rules. ^17^ Proteomic measures were complete and were not imputed.

Candidate mediators were identified in two screening steps. First, each exposure was regressed on every aptamer (**Figure 2**, Path A), and each outcome was regressed on every aptamer (**Figure 2**, Path B), in an unadjusted model and a model adjusted for the covariates described below. Within each regression family, P values were corrected for multiple testing using the Benjamini-Hochberg false discovery rate (FDR). ^18^ A protein was retained as a candidate mediator for a given exposure-outcome pair only if it met FDR significance on both path a and path b in the adjusted model, which is a necessary condition for mediation. This intersection step served as the primary dimension reduction.

**Figure 2.**
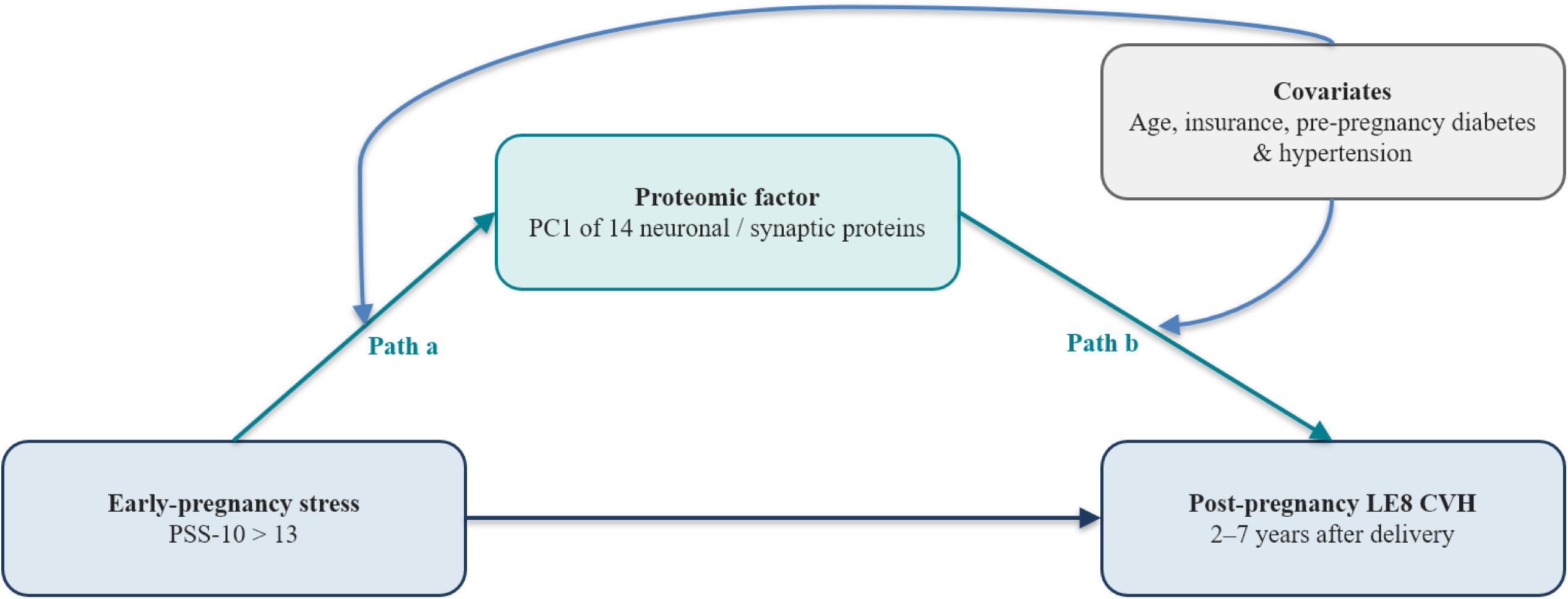
Analytic framework and proteomic screening. Schematic of the two-step screening in which early-pregnancy psychosocial exposures and post-pregnancy Life’s Essential 8 outcomes were each regressed on 6874 aptamers with Benjamini-Hochberg false discovery rate correction, and the intersection of significant proteins on both paths defined candidate mediators. Seventeen aptamers (14 unique proteins) met criteria for stress and the composite Life’s Essential 8 composite score and four clinical component scores.

All regression models were adjusted for maternal age, health insurance type (private, public, or other), and pre-pregnancy diabetes and pre-pregnancy hypertension as indicators of pre-pregnancy cardiometabolic health. Health insurance has been identified as one of the most important individual-level social determinants of health among pregnant individuals. ^5^ These covariates were selected as potential confounders of the exposure-outcome and mediator-outcome associations (**Figure 2**).

Principal component analysis (PCA) on the correlation matrix was used to reduce dimensionality of identified proteomic variables. The number of components was determined by the Kaiser criterion (eigenvalue >1) and the scree plot. The retained component score was standardized. Aptamers targeting the same protein were averaged before the PCA.

Mediation was quantified using the four-way decomposition, which partitions the total effect of the exposure into a controlled direct effect, a reference interaction, a mediated interaction, and a pure indirect effect. ^19,20^ We adopted the four-way decomposition over traditional mediation approaches because it distinguishes exposure-mediator interaction from mediation rather than conflating the two. ^21^ Models were fit with the *med4way* command using linear models for the mediator and outcome, contrasting stress (present vs absent), with covariates fixed at reference values (age at the sample median of 27 years; private insurance; no pre-pregnancy diabetes or hypertension). ^20^ The proportion mediated was calculated as the pure indirect effect divided by the total effect and reported when the total effect was statistically significant. Analyses were performed in Stata 18.5 (StataCorp LLC). Statistical significance was interpreted at two-sided *P* < .05 or 95% CIs excluding the null.

## Results

### Participant Characteristics

Among 1662 participants, the mean (SD) age was 27.1 (5.5) years; 62.2% identified as non-Hispanic White, 15.3% as non-Hispanic Black, and 15.0% as Hispanic. In the first trimester, 41.6% screened positive for perceived stress (PSS-10 >13), 17.3% for depression (EPDS >9), and 21.8% for anxiety (STAI >20). Pre-pregnancy hypertension and diabetes were present in 2.0% and 1.5%, respectively. The mean (SD) composite LE8 score was 78.7 (13.7) at the post-pregnancy visit. Distributions were similar before and after imputation (**Table 1** and **Table 2**). Sample characteristics were similar compared with all HHS participants and those without proteomics (**eTable 1**).

**Table 1.** Participant Characteristics in Early Pregnancy, Before and After Multiple Imputation.

|  | **Original**<br>**N=1,662** | **Imputed**<br>**(10 imputations)** | **p-value** |
| --- | --- | --- | --- |
| **Maternal age, years** | 27.1 (5.5) | 27.1 (5.5) | 1.00 |
| **Self-reported maternal race and ethnicity** |  |  | 1.00 |
| **NH White** | 62.2% | 62.2% |  |
| **NH Black** | 15.3% | 15.3% |  |
| **Hispanic** | 15.0% | 15.0% |  |
| **Asian** | 2.6% | 2.6% |  |
| **Other** | 5.0% | 5.0% |  |
| **Education attainment** |  |  | 1.00 |
| **High school or less** | 19.3% | 19.3% |  |
| **Some college** | 29.6% | 29.6% |  |
| **College or above** | 51.2% | 51.2% |  |
| **Health insurance at pregnancy** |  |  | 0.98 |
| **Others** | 3.1% | 3.1% |  |
| **Covered by public insurance** | 26.8% | 27.0% |  |
| **Covered by private insurance** | 70.0% | 69.8% |  |
| **Edinburgh Postnatal Depression Scale (0-30)** | 5.6 (4.2) | 5.6 (4.2) | 0.99 |
| **Depression (EPDS>9)** | 17.3% | 17.3% | 0.97 |
| **PSS-10 stress scale (0-40)** | 12.7 (6.8) | 12.7 (6.8) | 0.98 |
| **Stress (PSS-10>13)** | 41.6% | 41.6% | 0.98 |
| **State-Trait Anxiety Inventory (0-60)** | 13.8 (8.8) | 14.0 (8.8) | 0.52 |
| **Anxiety (STAI>20)** | 21.2% | 21.8% | 0.57 |
| **Fetal sex** |  |  | 0.99 |
| **Female** | 46.3% | 46.4% |  |
| **Male** | 53.7% | 53.6% |  |
| **Pre-gestational hypertension** | 2.0% | 2.0% | 1.00 |
| **Pre-gestational diabetes** | 1.5% | 1.5% | 1.00 |

Abbreviations: NH, non-Hispanic. Values are mean (SD) or %. The imputed column reflects estimates averaged across 10 imputed datasets. P values compare observed with imputed distributions.

**Table 2.** Cardiovascular Health Factors and Life’s Essential 8 Component and Composite Scores 2 to 7 Years After Delivery, Before and After Multiple Imputation.

|  | **Original**<br>**N=1,662** | **Imputed**<br>**(10 imputations)** | **p-value** |
| --- | --- | --- | --- |
| **Systolic Blood Pressure (mmHg)** | 112.2 (11.3) | 112.2 (11.3) | 0.97 |
| **Diastolic Blood Pressure (mmHg)** | 72.8 (10.2) | 72.8 (10.2) | 0.96 |
| **Non-HDL-cholesterol (mg/dL)** | 128.3 (38.4) | 128.3 (38.4) | 0.95 |
| **Glucose (mg/dL)** | 92.8 (27.2) | 92.8 (27.1) | 0.99 |
| **BMI (kg/m<sup>2</sup>)** | 28.5 (7.9) | 28.5 (7.9) | 0.93 |
| **Blood Pressure Score** | 84.1 (25.9) | 84.1 (25.9) | 0.97 |
| **Lipid Score** | 77.4 (28.0) | 77.4 (28.0) | 0.99 |
| **Glucose Score** | 90.0 (23.8) | 89.8 (24.0) | 0.81 |
| **BMI Score** | 64.7 (36.4) | 64.8 (36.4) | 0.93 |
| **LE8 CVH Score** | 78.8 (14.3) | 78.7 (13.7) | 0.68 |

Abbreviations: BMI, body mass index; LE8, Life's Essential 8; CVH, cardiovascular health; non-HDL, non-high-density lipoprotein cholesterol. Values are mean (SD). The imputed column reflects estimates averaged across 10 imputed datasets. P values compare observed with imputed distributions.

### Associations of Psychosocial Exposures with Post-Pregnancy CVH

In adjusted models, stress was associated with a lower post-pregnancy composite LE8 score (− 3.82 [95% CI, −5.11 to −2.54]) and a lower BMI score (−7.57 [95% CI, −11.1 to −3.99]); associations with the blood pressure, lipid, and glucose scores were not statistically significant. Similarly, depression was associated with a lower composite score (−3.56 [95% CI, −5.20 to − 1.92]) and a lower BMI score (−7.29 [95% CI, −11.9 to −2.67]). Anxiety was associated with a lower composite score (−4.66 [95% CI, −6.28 to −3.05]) and a lower BMI score (−7.57 [95% CI, − 11.9 to −3.27]) (**Table 3**).

**Table 3.** Associations of Early-Pregnancy Psychosocial Exposures with Post-Pregnancy Cardiovascular Health Outcomes.

|  |  | LE8 CVH Score | Blood Pressure Score | Lipid Score | Glucose Score | BMI Score |
| --- | --- | --- | --- | --- | --- | --- |
|  |  | Coef. (95% CI) | Coef. (95% CI) | Coef. (95% CI) | Coef. (95% CI) | Coef. (95% CI) |
| **Depression (EPDS>10)** | Unadjusted | -5.82 (-7.57, -4.07)* | -1.10 (-4.42, 2.22) | 0.44 (-3.24, 4.11) | -2.67 (-5.82, 0.47) | -10.6 (-15.2, -5.91)* |
|  | Adjusted | -3.56 (-5.20, -1.92)* | -0.058 (-3.34, 3.22) | -0.43 (-4.13, 3.27) | -1.09 (-4.05, 1.87) | -7.29 (-11.9, -2.67)* |
|  |  | Coef. (95% CI) | Coef. (95% CI) | Coef. (95% CI) | Coef. (95% CI) | Coef. (95% CI) |
| **Stress (PSS-10>13)** | Unadjusted | -6.05 (-7.37, -4.72)* | -3.09 (-5.62, -0.56)* | 1.20 (-1.57, 3.96) | -2.66 (-5.04, -0.28)* | -10.8 (-14.3, -7.31)* |
|  | Adjusted | -3.82 (-5.11, -2.54)* | -2.53 (-5.08, 0.03) | -0.40 (-3.23, 2.43) | -1.79 (-4.06, 0.48) | -7.57 (-11.1, -3.99)* |
|  |  | Coef. (95% CI) | Coef. (95% CI) | Coef. (95% CI) | Coef. (95% CI) | Coef. (95% CI) |
| **Anxiety (STAI>37)** | Unadjusted | -6.33 (-8.04, -4.62)* | -3.57 (-6.86, -0.28)* | -1.45 (-4.90, 1.99) | -3.12 (-6.23, -0.01)* | -10.1 (-14.5, -5.78)* |
|  | Adjusted | -4.66 (-6.28, -3.05)* | -2.25 (-5.42, 0.92) | -2.10 (-5.57, 1.38) | -2.13 (-5.04, 0.78) | -7.57 (-11.9, -3.27)* |

Each cell presents coefficients (95% Confidence Interval) based on a linear regression model.
Adjusted models included maternal age, insurance, pre-pregnancy diabetes, and pre-pregnancy hypertension. \* P < .05.

### Proteomic Screening and Candidate Mediators

Of 6874 aptamers, 17 aptamers, representing 14 unique proteins, were associated with both early-pregnancy stress and the post-pregnancy composite LE8 score after FDR correction in the adjusted model. The exact same set of aptamers were associated with stress and all four LE8 clinical outcomes after FDR correction. No aptamers met joint-association criteria for depression or anxiety, and the behavioral CVH components yielded few or no qualifying proteins.

Subsequent analyses therefore focused on stress and the composite LE8 score and the four clinical cardiometabolic outcomes. Three proteins were each represented by 2 aptamers (WFIKKN2, UNC5D, and NTM), and the concordance of these technical replicates supported the robustness of the signal.

Stress was associated with lower levels of all 14 proteins (Path A coefficients, −0.20 to − 0.24 per SD), and higher levels of each protein were associated with a better composite LE8 score (Path B coefficients, 2.4 to 4.8), yielding a directionally coherent pattern in which stress predicted a lower protein signature and a lower protein signature predicted worse CVH (**Table 4**; aptamer-level estimates and protein annotation in **eTable 2** and **eTable 3**). Notably, the candidate proteins were overwhelmingly neuronal and synaptic in function, including neural cell-adhesion and axon-guidance molecules (NRXN3, NTM, LSAMP, LRRTM2, UNC5D, PTPRD), myelin and neurotrophic proteins (MAG, CNTFR, APLP1), and secretory and scavenger proteins (SCG3, SCARA5, COLEC12, WFIKKN2, HS6ST3).

**Table 4.**
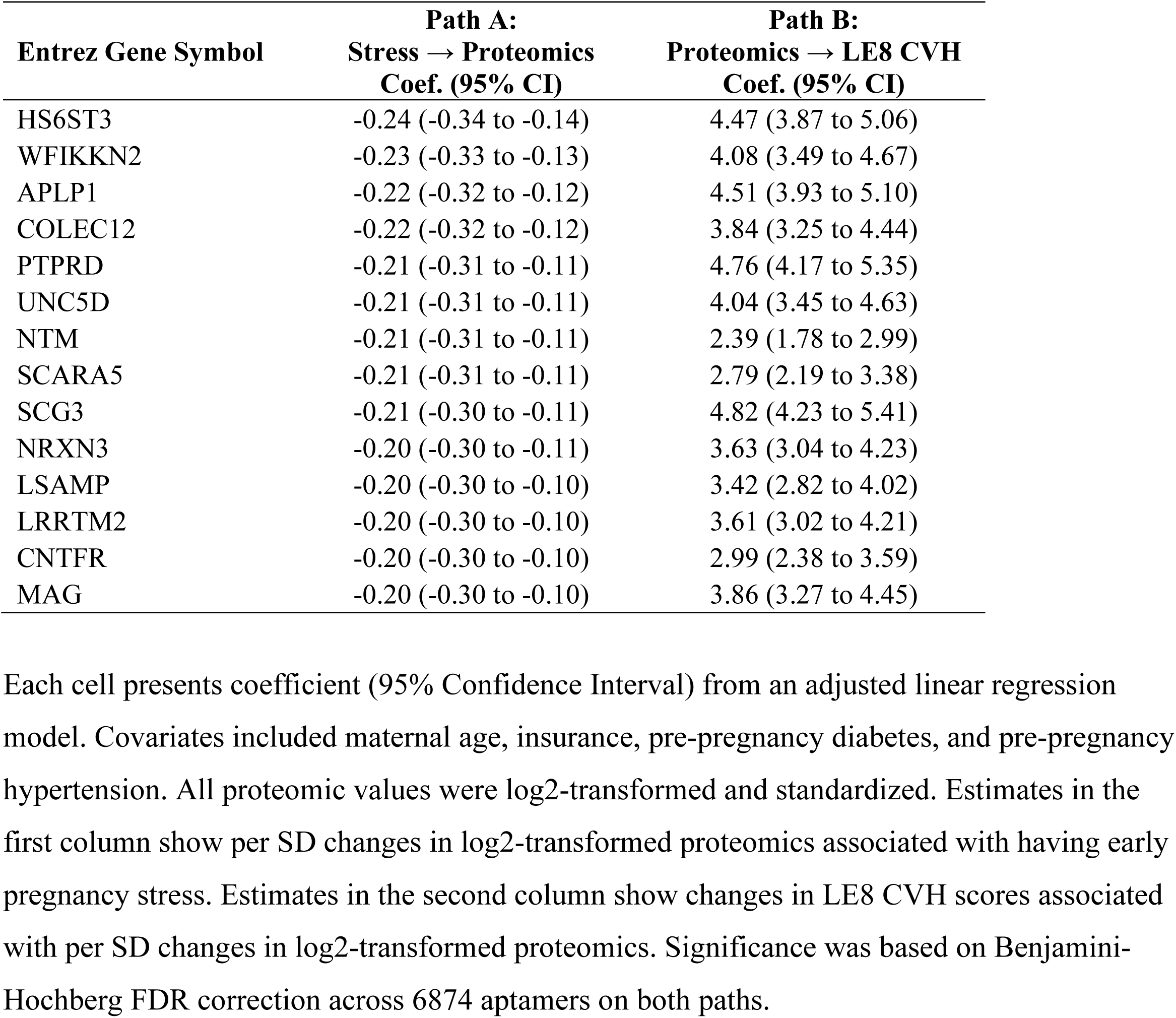
Joint Set of Proteins Associated with Both Early-Pregnancy Stress and Post-Pregnancy Life’s Essential 8 Outcomes.

The 14 proteins were highly intercorrelated and reduced to a single principal component, which explained 57% of the total variance (eigenvalue, 8.3); all subsequent eigenvalues were 1 or less. All 14 proteins loaded positively and uniformly on this component (loadings, 0.23-0.32), consistent with a single coherent protein signature (**eTable 4**). The standardized component score was used as the mediator in all mediation models.

### Mediation of Stress-CVH Associations by the Proteomic Factor

In the four-way decomposition, stress was associated with a lower post-pregnancy composite LE8 score (total effect, −3.74 [95% CI, −5.03 to −2.45]). The proteomic factor accounted for a pure indirect effect of −1.20 (95% CI, −1.68 to −0.72), approximately 32% of the total effect, with a controlled direct effect of −2.44 (95% CI, −3.64 to −1.25). For the BMI score, the total effect of stress was larger (−7.24 [95% CI, −10.87 to −3.62]) and the proteomic factor accounted for a pure indirect effect of −4.05 (95% CI, −5.61 to −2.49), approximately 56% of the total effect, together with a significant mediated interaction (−1.15 [95% CI, −2.10 to −0.20]) (**Table 5**).

**Table 5.** Four-Way Decomposition of the Association Between Early-Pregnancy Stress and Post-Pregnancy Cardiovascular Health, Mediated by the Standardized Proteomic Component.

|  | **LE8 CVH Score**<br>**Coef. (95% CI)** | **BP Score**<br>**Coef. (95% CI)** | **Lipid Score**<br>**Coef. (95% CI)** | **Glucose Score**<br>**Coef. (95% CI)** | **BMI Score**<br>**Coef. (95% CI)** |
| --- | --- | --- | --- | --- | --- |
| **Total effect** | -3.74 (-5.03 to -2.45) | -2.46 (-5.02 to 0.10) | -0.39 (-3.23 to 2.44) | -1.71 (-3.99 to 0.57) | -7.24 (-10.87 to -3.62) |
| **Controlled direct effect** | -2.44 (-3.64 to -1.25) | -1.21 (-3.74 to 1.33) | 0.96 (-1.85 to 3.77) | -0.48 (-2.72 to 1.77) | -2.81 (-6.01 to 0.39) |
| **Reference interaction** | 0.20 (-0.02 to 0.42) | 0.16 (-0.29 to 0.62) | 0.02 (-0.48 to 0.52) | 0.20 (-0.20 to 0.60) | 0.77 (0.13 to 1.41) |
| **Mediated interaction** | -0.30 (-0.63 to 0.03) | -0.25 (-0.92 to 0.43) | -0.03 (-0.78 to 0.72) | -0.29 (-0.89 to 0.31) | -1.15 (-2.10 to -0.20) |
| **Pure indirect effect** | -1.20 (-1.68 to -0.72) | -1.17 (-1.77 to -0.57) | -1.34 (-2.02 to -0.66) | -1.14 (-1.70 to -0.58) | -4.05 (-5.61 to -2.49) |

The total effect equals the sum of the controlled direct effect, reference interaction, mediated interaction, and pure indirect effect. Covariates were fixed at reference values (age 27 years; private insurance; no pre-pregnancy diabetes or hypertension). Unit of coefficients is LE8 score.

For the blood pressure, lipid, and glucose scores, the total effects of stress did not reach statistical significance, yet the pure indirect effects through the proteomic factor were statistically significant and tightly clustered (blood pressure, −1.17 [95% CI, −1.77 to −0.57]; lipids, −1.34 [95% CI, −2.02 to −0.66]; glucose, −1.14 [95% CI, −1.70 to −0.58]). For the lipid score, the significant adverse indirect effect was offset by a direct effect in the opposite direction (controlled direct effect, 0.96 [95% CI, −1.85 to 3.77]), producing a near-null total effect (**Table 5**).

## Discussion

In a prospective multicenter cohort of nulliparous individuals followed 2 to 7 years after a first delivery, early-pregnancy psychosocial stress was associated with worse post-pregnancy CVH, and a coherent signature of 14 predominantly neuronal and synaptic plasma proteins mediated approximately one-third of the association with the composite LE8 score and more than half of the association with BMI. Depression and anxiety did not yield qualifying protein mediators. To our knowledge, this is the first study to identify candidate circulating protein mediators linking maternal psychosocial stress to long-term cardiovascular health.

Three features of the proteomic signature strengthen its plausibility as a mediating pathway. First, the signature was directionally coherent. Stress was associated with lower protein levels, and lower protein levels were associated with worse CVH, across all 14 identified proteins. Second, the proteins were biologically unified. All identified proteins belong to neural cell-adhesion, axon-guidance, myelin, and neurotrophic molecules families, rather than a heterogeneous scatter, and collapsed cleanly to a single dimension. Third, the same factor produced consistent indirect effects across multiple cardiometabolic outcomes. Peripheral neurotrophic and synaptic-adhesion signaling has been implicated in the bidirectional communication between the brain and cardiometabolic systems, and stress-related neuroendocrine and inflammatory activation can plausibly downregulate such circulating proteins. ^22–24^ The findings suggest that the biological pathways connecting maternal stress to cardiovascular health may be partly captured by a measurable plasma protein signature in early pregnancy.

Notably, we observed significant indirect effects through the proteomic pathways for blood pressure, lipids, and glucose despite nonsignificant total effects. This pattern is expected when a real but modest mediated pathway is accompanied by limited power to detect the total effect, and, in the case of the lipid score, by a direct effect in the opposing direction that produces inconsistent mediation. The consistency of the indirect effects across cardiometabolic components (all approximately −1.2 points) reinforces the interpretation of a shared upstream biological pathway. The larger associations with BMI suggest that adiposity-related biology may be a particularly important node. The four-way decomposition proved advantageous over traditional mediation methods in our data. The association of stress with the BMI score included a significant mediated interaction that simpler decompositions would have absorbed into the indirect effect, indicating that stress and the proteomic factor act partly in concert rather than through mediation alone. Lastly, the specificity of the findings to stress, rather than depression or anxiety, may reflect the higher prevalence and statistical power for stress (41.6%) relative to depression (17.3%) anxiety (21.8%), differences in what each instrument captures, the timing of data collection for anxiety (mid-pregnancy), or a genuinely stronger biological link for perceived stress.

Several limitations warrant consideration. First, this was an observational analysis, and despite adjustment, residual and unmeasured confounding of the exposure-mediator, exposure-outcome, and mediator-outcome associations cannot be excluded. The decomposition estimates should be interpreted as quantifying pathways under standard mediation assumptions rather than established causal effects. Second, stress and plasma proteins were both measured in early pregnancy (although psychosocial questionnaires asked experience in time periods prior to data collection), so their temporal ordering cannot be firmly established and reverse influence of the proteome on perceived stress cannot be excluded. Third, proteins were measured on a single aptamer-based platform, and aptamer specificity and platform-specific coverage may affect which proteins were detected. Fourth, the proteomic subsample differed modestly from other HHS participants and the cohort comprised nulliparous individuals recruited largely at tertiary centers, which may limit generalizability.

In conclusion, in a prospective cohort of nulliparous individuals, a coherent neuronal and synaptic plasma protein signature in early pregnancy mediated a substantial portion of the association between maternal psychosocial stress and cardiovascular health 2 to 7 years after delivery. These findings nominate a measurable biological pathway that links maternal stress to long-term cardiovascular health and warrant replication on independent platforms and in cohorts with serial proteomic sampling.

## Article Information

Funding: This study received funding from the American Heart Association (24CDA1260466).

The nuMoM2b and nuMoM2b-HHS studies were supported by the Eunice Kennedy Shriver National Institute of Child Health and Human Development and the National Heart, Lung, and Blood Institute.

Role of the Funder/Sponsor: The funders had no role in the design and conduct of the study; collection, management, analysis, and interpretation of the data; preparation, review, or approval of the manuscript; or decision to submit the manuscript for publication.

Conflict of Interest Disclosures: None.

Data Sharing Statement: nuMoM2b and nuMoM2b-HHS data are available through the NICHD Data and Specimen Hub (DASH).

