## Supplemental Materials for "Plasma Proteomic Mediators of the Association Between Early-Pregnancy Psychosocial Stress and Cardiovascular Health 2 to 7 Years After Delivery: The nuMoM2b-HHS Study"

**eTable 1.** Characteristics of nuMoM2b-HHS Participants with and without First-Trimester Proteomics.

|  | **Total HHS1** | **No proteomics** | **Analytical sample** | **p-value** |
| --- | --- | --- | --- | --- |
|  | N=4,508 | N=2,846 | N=1,662 |  |
| **Maternal age, years** | 27.0 (5.6) | 27.0 (5.6) | 27.1 (5.5) | 0.59 |
| **Estimated gestational age at screening, weeks** | 11.4 (1.6) | 11.4 (1.6) | 11.5 (1.5) | 0.064 |
| **Gestational age at pregnancy end, weeks** | 38.7 (2.7) | 38.6 (3.0) | 38.8 (1.9) | 0.002 |
| **Time between index pregnancy and follow-up, years** | 3.2 (0.9) | 3.2 (0.9) | 3.2 (0.9) | 0.92 |
| **Self-reported maternal race and ethnicity** |  |  |  | 0.066 |
| **NH White** | 62.1% | 62.1% | 62.2% |  |
| **NH Black** | 13.8% | 13.0% | 15.3% |  |
| **Hispanic** | 16.3% | 17.1% | 15.0% |  |
| **Asian** | 3.0% | 3.2% | 2.6% |  |
| **Other** | 4.7% | 4.5% | 5.0% |  |
| **Nativity** |  |  |  | 0.94 |
| **US-born** | 87.3% | 87.3% | 87.4% |  |
| **Non-US-born** | 12.7% | 12.7% | 12.6% |  |
| **Education attainment** |  |  |  | 0.28 |
| **High school or less** | 18.6% | 18.3% | 19.3% |  |
| **Some college** | 31.0% | 31.8% | 29.6% |  |
| **College or above** | 50.4% | 49.9% | 51.2% |  |
| **Household income in 10K USD** | 8.3 (5.9) | 8.2 (5.9) | 8.6 (5.7) | 0.053 |
| **Health insurance at V1** |  |  |  | 0.64 |
| **Others** | 3.4% | 3.5% | 3.1% |  |
| **Covered by public insurance** | 27.3% | 27.6% | 26.8% |  |
| **Covered by private insurance** | 69.3% | 68.8% | 70.0% |  |
| **State-Trait Anxiety Inventory (0-60)** | 13.7 (8.7) | 13.7 (8.7) | 13.8 (8.8) | 0.57 |
| **Edinburgh Postnatal Depression Scale (0-30)** | 5.7 (4.2) | 5.7 (4.2) | 5.6 (4.2) | 0.44 |
| **Perceived Stress Scale-10 (0-40)** | 12.7 (6.7) | 12.7 (6.6) | 12.7 (6.8) | 0.90 |
| **Fetal sex** |  |  |  | 0.080 |
| **Female** | 48.0% | 49.1% | 46.3% |  |
| **Male** | 52.0% | 50.9% | 53.7% |  |
| **Pre-gestational diabetes** | 1.4% | 1.3% | 1.5% | 0.57 |
| **Pre-gestational hypertension** | 2.8% | 3.3% | 2.0% | 0.017 |

Abbreviations: NH, non-Hispanic. Values are mean (SD) or %. P values compare HHS1 participants with and without proteomic data.

**eTable 2.** Aptamer-Level Associations for the 17 Aptamers Meeting Joint-Association Criteria with Stress and Post-Pregnancy LE8.

| **Aptamer Seq ID** | **Entrez Gene Symbol** | **Path A:**  **Stress → Proteomics**  **Coef. (95% CI)** | **Path B:**  **Proteomics → LE8 CVH**  **Coef. (95% CI)** |
| --- | --- | --- | --- |
| seqid1889623 | HS6ST3 | -0.24 (-0.34 to -0.14) | 4.47 (3.87 to 5.06) |
| seqid1340823 | WFIKKN2 | -0.24 (-0.34 to -0.14) | 4.01 (3.42 to 4.61) |
| seqid323550 | WFIKKN2 | -0.23 (-0.33 to -0.13) | 4.15 (3.56 to 4.74) |
| seqid1630722 | UNC5D | -0.23 (-0.33 to -0.13) | 4.03 (3.43 to 4.62) |
| seqid721025 | APLP1 | -0.22 (-0.32 to -0.12) | 4.51 (3.93 to 5.10) |
| seqid54575 | COLEC12 | -0.22 (-0.32 to -0.12) | 3.84 (3.25 to 4.44) |
| seqid929615 | PTPRD | -0.21 (-0.31 to -0.11) | 4.76 (4.17 to 5.35) |
| seqid8428102 | NTM | -0.21 (-0.31 to -0.11) | 2.15 (1.55 to 2.76) |
| seqid104191 | SCARA5 | -0.21 (-0.31 to -0.11) | 2.79 (2.19 to 3.38) |
| seqid79572 | SCG3 | -0.21 (-0.30 to -0.11) | 4.82 (4.23 to 5.41) |
| seqid163238 | NRXN3 | -0.20 (-0.30 to -0.11) | 3.63 (3.04 to 4.23) |
| seqid29996 | LSAMP | -0.20 (-0.30 to -0.10) | 3.42 (2.82 to 4.02) |
| seqid2055038 | NTM | -0.20 (-0.30 to -0.10) | 2.62 (2.02 to 3.22) |
| seqid690414 | LRRTM2 | -0.20 (-0.30 to -0.10) | 3.61 (3.02 to 4.21) |
| seqid141012 | CNTFR | -0.20 (-0.30 to -0.10) | 2.99 (2.38 to 3.59) |
| seqid2057950 | MAG | -0.20 (-0.30 to -0.10) | 3.86 (3.27 to 4.45) |
| seqid514056 | UNC5D | -0.20 (-0.30 to -0.10) | 4.05 (3.46 to 4.64) |

Each cell presents coefficient (95% Confidence Interval) from an adjusted linear regression model. Covariates included maternal age, insurance, pre-pregnancy diabetes, and pre-pregnancy hypertension. All proteomic values were log2-transformed and standardized. Estimates in the first column show per SD changes in log2-transformed proteomics associated with having early pregnancy stress. Estimates in the second column show changes in LE8 CVH scores associated with per SD changes in log2-transformed proteomics. Significance was based on Benjamini-Hochberg FDR correction across 6874 aptamers on both paths.

**eTable 3.** Principal Component Loadings of the 14 Candidate Proteins on the Retained Component (PC1).

| **Entrez gene symbol** | **PC1 loading** |
| --- | --- |
| HS6ST3 | 0.24 |
| WFIKKN2 | 0.26 |
| APLP1 | 0.27 |
| COLEC12 | 0.25 |
| PTPRD | 0.32 |
| UNC5D | 0.31 |
| NTM | 0.24 |
| SCARA5 | 0.23 |
| SCG3 | 0.29 |
| NRXN3 | 0.28 |
| LSAMP | 0.26 |
| LRRTM2 | 0.30 |
| CNTFR | 0.23 |
| MAG | 0.25 |

**eTable 4.** Annotation of the 14 Identified Proteins.

| **Full name** | **Target name** | **Entrez gene symbol** | **UniProt identifier** |
| --- | --- | --- | --- |
| Heparan-sulfate 6-O-sulfotransferase 3 | H6ST3 | HS6ST3 | Q8IZP7 |
| WAP, Kazal, immunoglobulin, Kunitz and NTR domain-containing protein 2 | WFKN2 | WFIKKN2 | Q8TEU8 |
| Amyloid-like protein 1 | Amyloid-like protein 1 | APLP1 | P51693 |
| Collectin-12 | COLEC12 | COLEC12 | Q5KU26 |
| Receptor-type tyrosine-protein phosphatase delta | PTPRD | PTPRD | P23468 |
| Netrin receptor UNC5D | UNC5H4 | UNC5D | Q6UXZ4 |
| Neurotrimin | NTRI | NTM | Q9P121 |
| Scavenger receptor class A member 5 | SCAR5 | SCARA5 | Q6ZMJ2 |
| Secretogranin-3 | SCG3 | SCG3 | Q8WXD2 |
| Neurexin-3 | NRX3A | NRXN3 | Q9Y4C0 |
| Limbic system-associated membrane protein | LSAMP | LSAMP | Q13449 |
| Leucine-rich repeat transmembrane neuronal protein 2 | LRRT2 | LRRTM2 | O43300 |
| Ciliary neurotrophic factor receptor subunit alpha | CNTFR alpha | CNTFR | P26992 |
| Myelin-associated glycoprotein | MAG | MAG | P20916 |
